# The Effects of Cognitive Behavioral Therapy for Insomnia on Cardiovascular and Immunological Outcomes: A Randomized-Controlled Study

**DOI:** 10.64898/2026.06.30.26356933

**Authors:** Mathilde Reyt, Denise C. Jarrin, Aurore A. Perrault, Florence Borgetto, Dylan Smith, Kirsten Gong, Lukia Tarelli, Josée Savard, Thien Thanh Dang-Vu, Jean-Philippe Gouin

## Abstract

Evidence suggests that insomnia disorder is associated with pathophysiological alterations that may contribute to long-term physical, mental and inflammatory-related health risks. Cognitive behavioral therapy for insomnia (CBTi) is the first-line treatment for insomnia disorder, yet its effects on physiological outcomes remain unclear. This randomized-controlled trial examined the effects of CBTi on cardiovascular and immunological biomarkers.

Sixty-two participants with insomnia disorder were randomized to group-CBTi (*N* = 33, 75.8% female, *M_age_* = 48.8 ± 17.1 years) or Waitlist (WL) control (*N* = 29, 75.9% female, *M_age_* = 52.2 ± 15.6 years). Cardiovascular parameters included systolic blood pressure (SBP), diastolic blood pressure (DBP), heart rate (HR), and nocturnal heart rate variability (HRV). Inflammatory markers from blood samples included C-reactive protein (CRP), tumor necrosis factor-alpha (TNF- *α*), interleukin-6 (IL-6) and brain-derived neurotrophic factor (BDNF). All outcomes were assessed at baseline (T1), post-treatment assessment (T2, following completion of CBTi or WL period), and 6-months for the WL group (T3, after CBTi for the WL participants).

No significant Group-by-time effects were observed for SBP, DBP, HR, HRV and any inflammatory markers (*p_s_* > .05) from T1 to T2. When pooling treatment effects following CBTi exposure across both groups (T1 to T2 in CBTi group and T1 to T3 in WL group), no significant biomarker changes were observed. Overall, results indicate that CBTi did not produce detectable changes in cardiovascular or inflammatory markers among healthy individuals with insomnia disorder. These findings suggest physiological responses to CBTi are complex and may reflect dynamic and context-dependent processes (https://www.isrctn.com/ISRCTN13983243).

**HIGHLIGHTS:**

- Insomnia disorder is associated with physiological alterations
- We found no significant cardiovascular changes following CBTi
- No significant effects of CBTi on inflammatory biomarkers were observed
- The findings suggest dissociation between subjective and physiological outcomes of CBTi

## 1. Introduction

Insomnia disorder is characterized by nocturnal (e.g., sleep initiation and sleep maintenance difficulties, early morning awakenings) and diurnal complaints despite adequate opportunity for sleep [1]. Chronic insomnia is defined by symptoms occurring at least three times per week and persist for at least three months. Although insomnia is often conceptualized as a psychological disorder triggered by psychosocial factors and maintained by maladaptive behaviors and cognitions [2], evidence indicates that it also involves physiological dysregulation across cardiac, hormonal and immune systems [3]. This state of hyperarousal is considered a core feature of insomnia and an integrative framework explaining how psychological and physiological processes contribute to its development and persistence [3,4].

Accordingly, interventions targeting these mechanisms have been developed, among which cognitive behavioral therapy for insomnia (CBTi) is considered the first-line of treatment for the management of chronic insomnia. CBTi is a multimodal psychological intervention combining cognitive and behavioral strategies aiming at restructuring dysfunctional sleep beliefs and altering sleep-interfering behaviors and poor sleep hygiene [5]. Evidenced by numerous meta-analyses and systematic reviews of randomized controlled trials, CBTi demonstrates robust and durable reduction in insomnia severity across insomnia-only and comorbid populations (e.g., heart failure, psychiatric disorders) [6,7] and delivery modalities (e.g., face-to-face, group, digital) [8,9].

Insomnia is associated with physiological arousal, reflected by increased hypothalamic–pituitary–adrenal (HPA) axis activity (e.g., cortisol secretion) [10] and autonomic nervous system (ANS) activation (e.g., elevated body temperature, blood pressure (BP) and heart rate (HR)) [3,10], compared with participants without sleep complaints. More specifically, individuals diagnosed with insomnia disorder have been shown to exhibit reduced high frequency (HF) heart rate variability (HRV) both during wakefulness [11,12] and across all sleep stages [13]. However, studies have reported decreased or unchanged HPA axis activity, ANS activity and HRV in individuals with or without insomnia [11–17].

Recent studies have examined whether CBTi can modulate physiological alterations [18]. Across three studies, CBTi showed no significant effects on BP, including mean 24-h [19], day- and night-time [20], and resting BP [21]. Evidence for HR and HRV, is more limited and inconsistent. While some studies found no differences in mean 24-h, day- or night-time HR or HRV (e.g., standard deviation of normal-to-normal (NN) intervals (SDNN), the low frequency (LF) to HF ratio) between CBTi and control groups after treatment [20], another study reported dampened HRV during N2 and REM sleep following CBTi in patients with insomnia (without a control group); with no differences according to treatment response or remission status [22]. In contrast, responders to CBTi showed increased parasympathetic activity following intervention, with an increase in the proportion of successive NN interval differences greater than 50 ms, divided by the total number of NN intervals (pNN50) [23]. Similarly, a recent study observed increased HF and reduced LF/HF ratio among health care workers after a ‘single-shot’ of CBTi accompanied with an information pamphlet compared to the control group [24].

Hyperarousal in insomnia is also reflected in inflammatory biomarkers. Individuals with insomnia exhibit 24-h alterations in circadian rhythm, including changes in the pattern and (hyper)secretion of proinflammatory cytokines (e.g., increased tumor necrosis factor-alpha [TNF-*α*], interleukin-6 [IL-6] from mid-afternoon to evening) [25]. Insomnia has also been associated with elevated circulating inflammatory biomarkers (e.g., CRP, TNF-*α*, serum amyloid protein, granulocyte-macrophage colony-stimulating factor) [26,27]. Accordingly, reduced concentrations of BDNF and proBDNF, key regulators of neuroinflammation, have been reported in older adults with insomnia [28,29]. A meta-analysis study further confirmed these associations, showing higher CRP and IL-6 levels in relation to sleep disturbances, as well as elevated CRP in short sleep duration [30].

Research on the effect of insomnia interventions on inflammation have produced mixed results. Across intervention studies (including CBTi, Tai Chi Chih and sleep seminar education), CBTi has been associated with reductions in inflammatory markers (e.g., CRP, proinflammatory gene expression and cytokines) [31,32] and cardiovascular, metabolic and inflammatory disease risk, as well as higher remission rates at 4- and 16-month follow-ups, compared with control conditions [33]. Similarly, one study in hemodialysis patients reported decreases in markers of inflammatory and oxidative stress (including CRP) following CBTi compared with a control group [34]. However, null or mixed effects have also been reported: CRP and TNF-*α* levels did not significantly decrease following CBTi but were reduced following Tai Chi Chih in breast cancer survivors [35], and no post-treatment differences in proinflammatory cytokines (IL-6, serum-soluble tumor necrosis factor receptor 2) were observed between CBTi and psychoeducation in patients with insomnia and bipolar disorder. Several additional studies in diverse populations (e.g., college students with irritable bowel syndrome, insomnia, short sleep duration heart failure) have similarly found no significant changes in inflammatory biomarkers (notably, IL-6) following CBTi [7,20,36,37]. Only one study examined BDNF levels, reporting no significant effects of CBTi [38]. Overall, evidence remains inconsistent regarding the impact of CBTi on inflammatory biomarkers.

Insomnia is a highly prevalent disorder with substantial impact on health [2]. Accordingly, CBTi and its associations with physiological markers warrant further investigation. Specifically, whether CBTi can reduce current or future disease risk through underlying pathophysiological changes remains unclear. Previous studies have focused on single physiological systems or combined markers, often using active control conditions (e.g., sleep education), which may attenuate between-group differences through non-specific effects. Wait-list controls, in contrast, provide a clearer estimate of the intervention effect. The present study addresses these gaps by conducting a secondary analysis of a randomized controlled trial that previously demonstrated a significant reduction in insomnia severity following CBTi, compared with a waitlist control condition [39]. We simultaneously assessed cardiovascular and inflammatory biomarkers in adults with chronic insomnia. The primary aim was to determine whether CBTi would produce significant and clinically meaningful pre- to post-treatment changes in these biomarkers. We hypothesized that CBTi would reduce BP, HR, and inflammation, while increasing HRV parameters.

## 2. Materials and methods

### 2.1. Sample Context and Design

This study reports secondary analyses of data from a randomized-controlled trial investigating the effects of CBTi on sleep and cognitive function, using combined objective and subjective assessments [39] (project registered as clinical trial ISRCTN13983243). Information on the participants, procedures, and results concerning the original study and primary research aims have been published elsewhere [39]. Briefly, participants completed evaluations at baseline and were then randomized to receive treatment (TX group) or wait for treatment (waitlist [WL] control group). Participants receiving treatment were assessed at 3-months post-treatment. Participants in the WL condition were assessed 3 months after baseline (i.e., following the WL period) and at 6 months post-baseline, after completing CBTi. As part of the larger study, participants completed several assessments (e.g., battery of questionnaires, sleep diaries, neurocognitive tasks, polysomnography [PSG], collection of physiological data); however, only variables pertinent to the present study will be described in detail.

### 2.2. Participants

Participants met DSM-5 criteria for chronic insomnia, confirmed through a semi-structured clinical interview. Participants with other sleep disorders were excluded (apnea-hypopnea index >15 events/h; periodic limb movement index >15 events/h). Additional details regarding recruitment, eligibility criteria, study procedures and the CONSORT flow diagram have been published previously [39]. In total, 62 participants were formally enrolled in the study.

Written informed consent was obtained before the start of the study. The study was approved by the Concordia University Human Research Ethics Committee and conducted in accordance with institutional ethical guidelines, ensuring respect for participants’ privacy rights.

### 2.3. Procedure

Approximately one month after the initial PSG screening, participants completed baseline assessments (T1), including a second PSG (baseline), questionnaires, morning ambulatory cardiovascular recordings, and blood draws. Questionnaires were completed on the night of the second PSG recording, while BP and HR were recorded the following morning. Blood draws were collected on the morning of the overnight study. Participants were then randomized to a 3-month CBTi treatment program (*n*=33) or a 3-month WL control condition (*n*=29). At 3 months post-randomization (T2), all participants repeated baseline assessments. The WL group subsequently received the CBTi program and completed a final assessment 3 months later (T3) (i.e., 6-month post-randomization).

### 2.4. Study Conditions

#### 2.4.1. CBTi Treatment

The CBTi program was delivered by a licensed psychologist over eight group sessions (2–6 participants per each group; *n*=14 groups). Sessions were provided over 3 months, including weekly meetings for 6 weeks followed by two sessions at 2–3 weeks intervals (1.5h each). The intervention included cognitive, behavioral, relaxation and educational components [5]. Cognitive restructuring aimed to alter misconceptions and maladaptive beliefs about sleep (e.g., unrealistic sleep expectations). Behavioral strategies included sleep restriction and stimulus control. The educational component addressed sleep physiology, circadian rhythms, and sleep hygiene (e.g., caffeine use, exercise, environmental factors). Relaxation training included progressive muscle relaxation and deep breathing techniques.

#### 2.4.2. Wait-List (WL) Condition

The WL control condition consisted of a 3-month monitoring period. A 3-month waiting period is equal to or shorter than what a patient typically waits for to obtain treatment in the public health care system, and thus, was considered ethically appropriate. CBTi treatment was provided to the WL participants following the 3-month waiting period.

### 2.5. Ambulatory Cardiovascular Measures

#### 2.5.1. Resting BP and HR

Participants underwent a 5-minute resting-state recording in the morning, during which they were instructed to close their eyes. During this period, beat-to-beat BP was measured using a Continuous Noninvasive Arterial Pressure monitor with arm and finger cuffs. Continuous ECG (electrocardiogram) recordings (further described below) were obtained to derive resting HR. All data were visually inspected for artifacts or outliers; three measures for BP and HR were taken and averaged.

#### 2.5.2. Nocturnal HRV

Continuous ECG recordings were used to assess nocturnal HRV using a standard Lead II configuration with three pre-gelled Ag/AgCl electrodes positioned at the jugular notch of the sternum, approximately 4 cm below the left nipple, and on the right lateral thoracic wall. Signals were acquired with the MindWare BioNex system at 1000 Hz. R-peaks were detected using MindWare HRV software (version 3.1.5; Mindware Technologies), and the resulting inter-beat intervals (IBI) were exported for offline processing in Python using the hrvanalysis package (version 1.0.5). RR intervals outside physiological ranges (<300 ms or >1800ms) were removed, and ectopic beats were corrected using the Malik method [40]. Missing intervals were linearly interpolated to obtain a continuous IBI time series. All corrected tachograms were visually inspected prior to HRV analyses to ensure data quality.

Following preprocessing, HRV analyses were conducted using Kubios HRV Analysis software (Kubios HRV Scientific 4.2.0, Kuopio, Finland) during two sleep intervals: the first 90 minutes from sleep onset and the full overnight period (from sleep onset to final awakening). Time-domain measures included SDNN, reflecting overall autonomic variability, and the root mean square of successive differences (RMSSD), an index primarily reflecting parasympathetic modulation of heart rate. Frequency-domain indices were derived from power spectral density estimation using Fast Fourier Transform and included low frequency (LF: 0.04–0.15 Hz) and high frequency (HF: 0.15–0.42 Hz), with HF power indexes parasympathetic activity whereas LF reflects mixed sympathetic-parasympathetic influences. LF and HF values were log-transformed prior to statistical analyses to normalize their distributions.

### 2.6. Inflammatory Markers

Blood samples were obtained via antecubital venipuncture in the morning following each sleep assessment (e.g., T1, T2, T3), between 7h00 to 11h00 after an overnight fast. Samples were collected in EDTA-coated vacutainer tubes, centrifuged, aliquoted, and stored at −80°C until analysis. Circulating inflammatory and neurotrophic biomarkers including CRP, TNF-*α*, IL-6 and BDNF were assayed in duplicate using multiplex V-Plex kits (MSD, Meso Scale Discovery, Rockville, MD). Due to positive skewness, values were log10-transformed to normalize their distributions.

### 2.7. Statistical Analysis

All analyses were performed using R version 4.4.1 and R packages (e.g., tidyverse, lme4, lmerTest, emmeans and effectsize). To examine the effects of CBTi (TX group) compared to a waitlist control (WL group) on inflammatory and cardiovascular markers, linear mixed-effects models were conducted for each outcome variable separately. Each model included Group (TX vs. WL), Time (Pre vs. Post intervention), and their interaction (Group × Time) as fixed effects, with age and sex included as covariates. A random intercept for each subject was included to account for repeated measures within individuals. Outliers were identified based on standardized residuals from the main model. Observations with absolute standardized residuals greater than 3 were considered outliers and excluded from the analyses. Significance of fixed effects was tested using Type III analyses of variance (ANOVA). The primary effect size measure was partial omega squared (ω²p) for the Group × Time interaction, reflecting the proportion of variance attributable to the treatment effect over time. Values of 0.01, 0.06, and 0.14 were interpreted as small, medium, and large effects, respectively. Data were analyzed with an intention-to-treat approach. Linear mixed-effects models include all randomized participants and make use of all available data under a maximum likelihood framework, without requiring explicit imputation of missing data, assuming data are missing at random. Graphical diagnostics and Shapiro-Wilk test were used to check the normality of the distribution of each outcome variable (BP, HR, HRV and inflammatory markers). Levene’s test was used to examine homogeneity of variance.

To further examine the effects of CBTi exposure over time and increase statistical power, we pooled together T2 data from participants who initially received CBTi and T3 data from participants who were initially assigned to the waitlist condition and subsequently received CBTi. Linear mixed-effects models were used for each outcome, including Time as a fixed effect, and age and sex as covariates. A random intercept for each subject was specified to account for repeated measures within individuals. Type III ANOVA was performed on each model to evaluate the significance of the main effect. This approach allowed us to assess changes over time while adjusting for individual differences and covariates.

For comparisons of demographic variables between groups, statistical significance was defined as *p*<.05. Sex was defined as self-reported sex assigned at birth and recorded as a biological variable in the study. The Benjamini-Hochberg False Discovery Rate (FDR) procedure was applied separately to each family of related outcomes (cardiovascular, inflammatory and HRV analyses) to account for multiple testing.

## 3. Results

### 3.1. Demographics

From the 62 participants included, 33 (75.8% female; *M*=48.8 years, *SD*=17.1, range: 22 to 82 years, mean insomnia duration =12.41 years, *SD*=15.85) were assigned into the TX condition. Twenty-nine participants were randomized into the WL condition (75.9% female; *M*=52.2 years, *SD*=15.6, range: 21 to 74 years, mean insomnia duration =12.55 years, *SD*=12.12). A majority of the participants were White (74.2%), female (75.8%), completed university (69.4%), non-smokers (91.4%), had optimal BP readings (<120/<80 mmHG; 83.3%), and did not have any major psychological disorders other than insomnia (82.3%). No sex (*X^2^*<0.001, *df*=1, *p*=0.99) or age differences (Welch t-test, *t*=-0.81, *df*=59.91, *p*=0.42) between the two groups were found.

In the TX condition, 30 of 33 participants (90.9%) completed the T2 assessment. No age or sex differences were observed between participants who completed assessments from T1 to T2 in the TX condition (*p_s_*>.05). In the WL condition, 23 of 29 participants (79.3%) completed the T2 assessment, and 22 of 29 participants (75.9%) completed the T3 assessment. No age or sex differences were observed between participants who completed assessments from T1 to T2 in the WL condition (*p_s_* > .05). However, participants who did not complete T3 assessments (*M*=39 years, *SD*=18.09) were significantly younger than those that did complete assessments (*M*=56.4 years, *SD*=12.39) with a trend toward significance (*t*=2.27, *df*=7.88, *p*=.05). No significant sex differences were observed between participants who completed assessments from T1 to T3 in the WL condition (*p*>0.05). Detailed information on participant retention and missing data for each outcome measure is provided in the Supplementary Methods.

### 3.2. Morning Ambulatory Cardiovascular Measures

Table 1 presents descriptive statistics for SBP, DBP, HR, and HRV indices. Hegde’s *g* values are reported in the Supplementary Results (Table S1). No significant between-group differences were found at baseline across these variables, although DBP showed a trend toward group differences (*p*=0.05). A linear mixed-effects model was conducted to examine the effects of the CBTi intervention on SBP, controlling for age and sex. The fixed effects revealed no significant Group x Time interaction (*F*(1, 49.49)=0.57, *p*=0.45, *p*_FDR_=0.68, ω²p<.01). There was no main effect of Time (*F*(1, 49.17)=0.01, *p*=0.94, *p_FDR_*=0.94), nor significant main effect of Group either (*F*(1, 56.67)=2.36, *p*=0.13, *p_FDR_*=0.33).

**Table 1.**
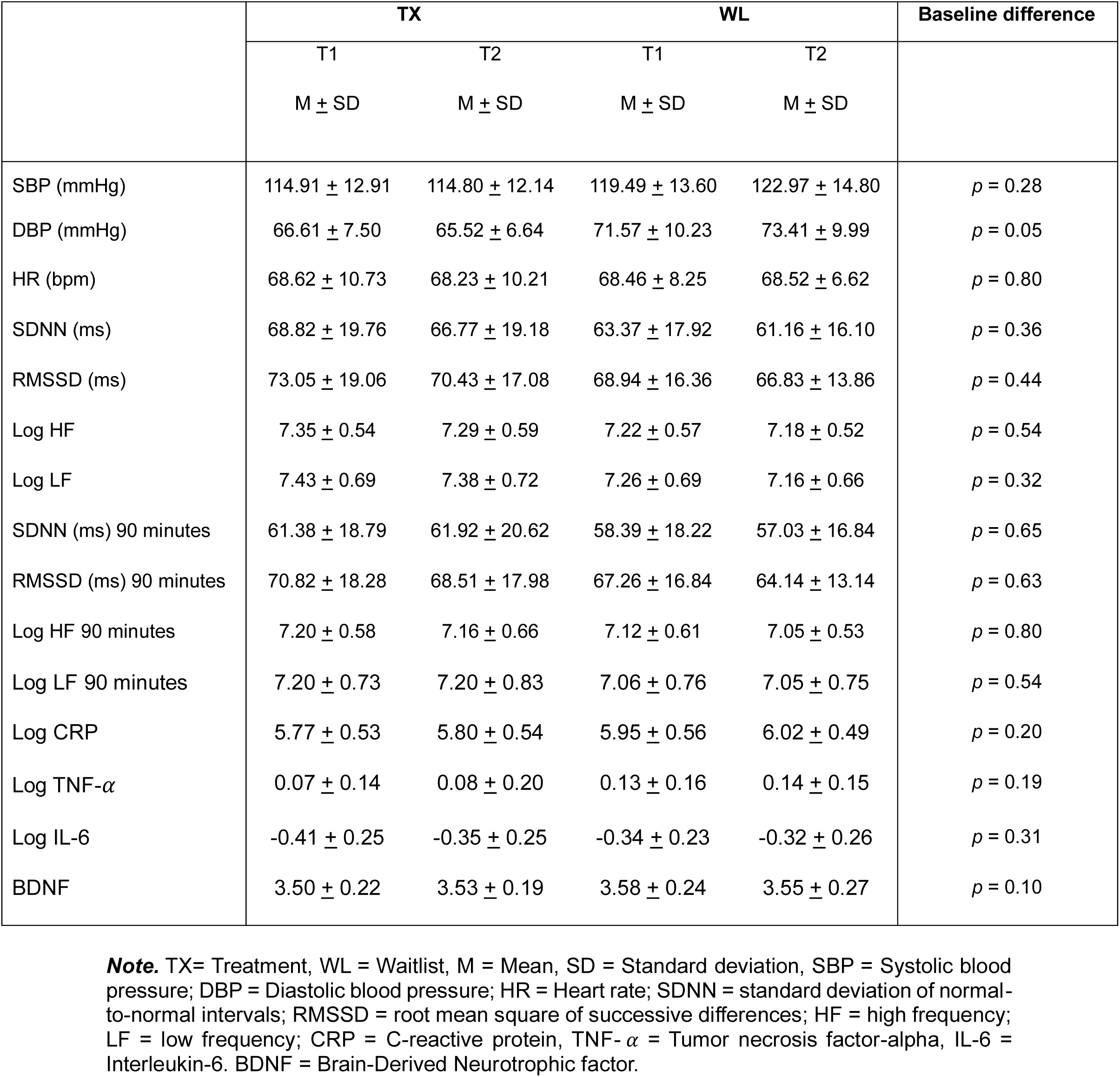
Descriptive Statistics for Cardiovascular and Immunological Measures at T1 and T2.

No significant Time (*F*(1, 50.01)=1.16, *p*=0.29, *p_FDR_*=0.49) or Group x Time interaction effects (*F*(1, 50.14)=0.25, *p*=0.62, *p_FDR_*=0.76, ω²p<.01) were found for DBP. A difference between groups was observed for DBP with an uncorrected *p*-value (*F*(1, 57.71)=5.51, *p*<.05), but this difference did not remain significant after FDR correction (*p_FDR_*=0.09).

There were no significant effects of Time (*F*(1, 50.64)=0.22, *p*=0.64, *p_FDR_*=0.76), Group (*F*(1, 58.47)=0.17, *p*=0.68, *p_FDR_*=0.76), or their interaction (*F*(1, 50.82)=1.11, *p*=0.30, *p_FDR_*=0.49, ω²p< .01) on HR.

We then examined the treatment effects following CBTi exposure in both groups pooled together, with changes observed in the TX group (T1 to T2) and in the WL group after receiving CBTi (T1 to T3). Descriptive statistics for the pooled data are presented in Table 2. Corresponding effect sizes are reported in the Supplementary Results (Table S2).

No main effect of Time was observed for SBP (*F*(1, 50.01)=1.85, *p*=0.18, *p_FDR_*=0.32), DBP (*F*(1, 49.32)=1.38, *p*=0.25, *p_FDR_*=0.32]) and HR (*F*(1, 47.59)=0.01, *p*=0.99, *p_FDR_*=0.99).

### 3.3. Nocturnal Ambulatory Cardiovascular Measures

Descriptive statistics for time- and domain-specific frequency during sleep and for the first 90 minutes of sleep are presented in Table 1. Effect size (Hegde’s *g*) are reported in the Supplementary Results (Table S1). At baseline, no significant differences were observed between groups (*p_s_*>0.05, Table 1). No significant effect of Group (*F*(1, 55.06)=1.17, *p*=0.28, *p_FDR_*=0.59), nor Time (*F*(1, 41.94)=0.63, *p*=0.43, *p_FDR_*=0.62), or their interaction (*F*(1, 41.92)=0.17, *p*=0.68, *p_FDR_*=0.78, ω²p<.01) were observed on SDNN during the whole night. Similar results were found on SDNN during the first 90 minutes of sleep (Time: *F*(1, 43.89)=0.11, *p*=0.74, *p_FDR_*=0.84; Group: *F*(1, 55.52)=0.33, *p*=0.57, *p_FDR_*=0.83; Group x Time: *F*(1, 43.86)=0.18, *p*=0.67, *p_FDR_*=0.84, ω²p<.01).

No significant effects were observed for RMSSD during sleep (Time: *F*(1, 42.86)=0.95, *p*=0.34, *p_FDR_*=0.60; Group: *F*(1, 55.45)=0.59, *p*=0.45, *p_FDR_*=0.62; Group x Time: *F*(1, 42.76)=0.006, *p*=0.94, *p_FDR_*=0.96, ω²p<.01). Likewise, RMSSD remained unchanged over time and between groups during the first 90 minutes of sleep (Time: *F*(1, 42.80)=1.07, *p*=0.31, *p_FDR_*=0.64; Group: *F*(1, 54.65)=0.53, *p*=0.47, *p_FDR_*=0.83; Group x Time: *F*(1, 42.78)=0.24, *p*=0.63, *p_FDR_*=0.84, ω²p<.01).

Analyses revealed no significant effects for HF during sleep (Time: *F*(1, 43.75)=0.49, *p*=0.49, *p_FDR_*=0.64; Group: *F*(1, 55.47)=0.38, *p*=0.54, *p_FDR_*=0.68; Group x Time: *F*(1, 43.73)=0.10, *p*=0.76, *p_FDR_*=0.82, ω²p<.01). Restricting the analyses to the first 90 minutes of sleep yielded comparable findings (Time: *F*(1, 43.45)=0.71, *p*=0.41, *p_FDR_*=0.78; Group: *F*(1, 55.35)=0.15, *p*=0.70, *p_FDR_*=0.84; Group x Time: *F*(1, 43.43)=0.05, *p*=0.82, *p_FDR_*=0.85, ω²p<.01).

Similarly, LF remained stable across conditions (Time: *F*(1, 42.45)=0.60, *p*=0.44, *p_FDR_*=0.62; Group: *F*(1, 55.37)=1.07, *p*=0.31, *p_FDR_*=0.59; Group x Time: *F*(1, 42.37)=0.27, *p*=0.60, *p_FDR_*=0.72, ω²p<.01). A similar pattern was observed during the first 90 minutes of sleep (Time: *F*(1, 44.43)=0.13, *p*=0.72, *p_FDR_*=0.84; Group: *F*(1, 56.74)=0.39, *p*=0.54, *p_FDR_*=0.83; Group x Time: *F*(1, 44.32)=0.02, *p*=0.88, *p_FDR_*=0.88, ω²p<.01).

When data from both groups were combined (Table 2 and S2), analyses revealed no significant variations of SDNN over time during the first 90 minutes of sleep (*F*(1, 47.98)=0.05, *p*=0.82, *p_FDR_*=0.89) and also during the whole night (*F*(1, 46.32)=0.01, *p*=0.91, *p_FDR_*=0.98). No statistically significant differences were found for RMSSD during the whole night (*F*(1, 44.87)=0.14, *p*=0.71, *p_FDR_*=0.86), with comparable results for the initial 90 minutes of sleep (*F*(1, 43.84)=0.09, *p*=0.76, *p_FDR_*=0.89). No significant effect of Time was observed for frequency-domain measures, either across the whole night (HF: *F*(1, 46.01)=0.17, *p*=0.69, *p_FDR_*=0.86; LF: *F*(1, 46.51)=0.01, *p*=0.98, *p_FDR_*=0.98) or during the first 90 minutes of sleep (HF: *F*(1, 46.15)=0.40, *p*=0.53, *p_FDR_*=0.71; LF: *F*(1, 48.13)=0.003, *p*=0.96, *p_FDR_*=0.96).

### 3.3. Inflammatory Markers

Descriptive statistics for inflammatory markers are found in Table 1 and the corresponding effects sizes in Table S1. No significant effects were observed for CRP (Time: *F*(1, 49.75)=0.03, *p*=0.87, *p_FDR_*=0.92; Group: *F*(1, 57.15)=2.14, *p*=0.15, *p_FDR_*=0.63; Group x Time: *F*(1, 49.83)=0.09, *p*=0.76, *p_FDR_*=0.92, ω²p<.01) or for TNF-*α* (Group: *F*(1, 57.18)=2.05, *p*=0.16, *p_FDR_*=0.63; Time: *F*(1, 49.29)=0.10, *p*=0.75, *p_FDR_*=0.92; Group x Time: *F*(1, 49.42)=0.24, *p*=0.63, *p_FDR_*=0.92, ω²p<.01). Similarly, no significant effects were observed for IL-6 (Time: *F*(1, 49.96)=0.35, *p*=0.56, *p_FDR_*=0.92; Group: *F*(1, 56.54)=0.62, *p*=0.44, *p_FDR_*=0.91; Group x Time: *F*(1, 50.11)=0.42, *p*=0.52, *p_FDR_*=0.92, ω²p<.01). In line with these results, analyses revealed no significant effects for BDNF (Group: *F*(1, 58.33)=0.92, *p*=0.34, *p_FDR_*=0.86; Time: *F*(1, 51.79)=0.001, *p*=0.98, *p_FDR_*=0.98; Group x Time: *F*(1, 51.93)=1.07, *p*=0.31, *p_FDR_*=0.86, ω²p<.01).

When data were pooled across all participants (Table 2 and S2), analyses revealed non-significant intervention effects between pre- and post-treatment for CRP (*F*(1, 49.07)=0.05, *p*=0.83, *p_FDR_*=0.83), TNF-*α* (*F*(1, 47.61)=0.85, *p*=0.36, *p_FDR_*=0.72) or IL-6 (*F*(1, 49.69)=2.61, *p*=0.11, *p_FDR_*=0.34). Similarly, no main effect of Time was observed for BDNF (*F*(1, 51.63)=0.87, *p*=0.35, *p_FDR_*=0.72).

### 3.4. Effects of covariates: age and sex

Age and sex were included as covariates in linear mixed-effects models. Significant age-related effects were observed across several cardiovascular measures. Specifically, SBP (*F*(1, 59.44)=15.81, *p*<.001, *p_FDR_*<.01) and DBP (*F*(1, 59.10)=8.19, *p*<.01, *p_FDR_*<.05) increased significantly with age. Age was significantly associated with RMSSD across the whole night and during the first 90 minutes of sleep: however, this association did not remain significant after FDR correction (whole night: *F*(1, 56.42)=4.28, *p*<.05, *p_FDR_*=0.36; 90 minutes: *F*(1, 55.21)=4.05, *p*<.05, *p_FDR_*=0.31). A similar trend was observed for HF within the first 90 minutes of sleep (*F*(1, 55.88)=4.24, *p*<.05, *p_FDR_*=0.31). When data were pooled across all participants to assess treatment effects following CBTi exposure, the age-related effect remained significant for SBP (*F*(1, 59.89)=16.89, *p*<.001, *p_FDR_*<.01) and DBP *F*(1, 60.56)=7.99, *p*<.01, *p_FDR_*<.05).

Although initial analyses suggested sex-related differences for HR (*F*(1, 56.56)=5.61, *p*<.05, *p_FDR_*=0.09), SBP (*F*(1, 58.81)=4.12, *p*<.05, *p_FDR_*=0.11), RMSSD (whole night: *F*(1, 56.50)=4.55, *p*<.05, *p_FDR_*=0.16; 90 minutes: *F*(1, 55.78)=4.51, *p*< .05, *p_FDR_*=0.15) and HF (whole night: *F*(1, 57.54)=5.33, *p*<.05, *p_FDR_*=0.16; 90 minutes: *F*(1, 57.65)=5.33, *p*<.05, *p_FDR_*=0.15), these associations did not withstand FDR correction. However, when data were pooled across all participants to assess treatment effects following CBTi exposure, a significant sex effect on HR was found, with females exhibiting higher HR than males (*F*(1, 58.87)=7.69, *p*<.01, *p_FDR_*<.05).

## 4. Discussion

Using a randomized-controlled design, the present study aimed to investigate the effects of CBTi, compared to a WL control condition, on cardiovascular and inflammatory markers among individuals with chronic insomnia. Alongside the substantial improvements in insomnia severity previously reported in this sample [39], CBTi intervention did not seem to improve cardiovascular or inflammatory markers.

### 4.1. Cardiovascular outcomes following CBTi

Although insomnia has been associated with a range of adverse health outcomes, the evidence remains heterogeneous, highlighting the need for additional studies to strengthen the current evidence base [41]. Therefore, randomized controlled trials evaluating the effects of CBTi on physiological markers of cardiovascular risk are needed to clarify whether improving insomnia can causally influence cardiovascular health.

In the present study, CBTi did not yield significant effects on BP, HR and HRV parameters. These results are consistent with the existing literature on the effects of CBTi on BP [19]. It is possible that a 3-month follow-up interval used in the present study, although longer than previous studies, may have still been insufficient to demonstrate potential benefits of CBTi on cardiovascular measures. Longer follow-up periods may therefore be required, as previously suggested in a systematic review of CBTi trials reporting cardiovascular markers [18]. Floor and ceiling effects could have also prevented the documentation of cardiovascular improvements following intervention. Specifically, most of the participants in the present study had normal BP readings across all assessments (mean T1: 83.3%; T2: 88.2%; T3: 87.5%), leaving limited opportunities for improvements. It is therefore possible that inclusion of higher-risk participants would have yielded more pronounced clinical effects. This interpretation is consistent with previous CBTi trials, where no effects on BP were observed in samples without cardiovascular disease [19,20] whereas small reductions in SBP (Cohen’s *d*=0.22) and DBP (Cohen’s *d*=0.25) were reported in an older sample with coronary heart disease following web-based CBTi [21]. Together, these findings suggests that baseline cardiovascular risk may moderate the effect of CBTi on BP outcomes.

The present study found no significant changes for diurnal HR from pre- to post-CBTi, aligning with findings from two studies that investigated HR in the morning [23] and over 24-h [20]. Significant effects on cardiometabolic measures have mainly been documented in vulnerable samples. Following CBTi and behavioral sleep intervention (based on CBTi), reduced insulin resistance among pregnant women [42] and improved glycemic control among insomnia patients with type 2 diabetes [43] have been documented. Following CBT interventions targeting other conditions, improvements in cardiometabolic measures have also been reported in populations with posttraumatic stress disorders [44,45]. Although this study did not observe significant change in cardiovascular risk, CBTi may have greater benefits in susceptible or clinically affected populations.

Our results did not show any significant effect of CBTi on HRV parameters, suggesting that the intervention may not induce detectable changes in cardiac autonomic regulation when HRV is evaluated over the entire night of sleep. The limited and heterogenous nature of the current literature in CBTi interventions on autonomic markers were conducted under markedly different methodological conditions. Two studies focused on short recordings (5 minutes) during wakefulness [23,24], which may capture transient autonomic responses but do not necessarily reflect sustained nocturnal autonomic regulation. One study restricted HRV analyses to specific sleep stages, with evidence suggesting a reduction in HF during N2 and REM sleep [22]. In contrast, our analysis considered HRV across the entire night, which may have diluted potential stage-specific effects of CBTi on autonomic function. This is in line with the growing view that brain activity during sleep actively contributes to fluctuations in cardiac autonomic regulation [46], emphasizing the importance of accounting for sleep-stage dynamics when assessing HRV responses to therapeutic interventions. Furthermore, stage-specific examination of HRV may help explain why some studies report autonomic differences between good sleepers and insomnia patients [13,15] whereas others do not [11,14,16].

### 4.2. Inflammatory responses to CBTi

Findings on the effects of CBTi on inflammatory markers remain inconsistent across studies and this heterogeneity is reflected in the lack of significant effects observed in the present study. These results are consistent with much of the research that found no effects on CRP [20,35,36], TNF-*α* [35,47], IL-6 [35,36] and BDNF [38] following CBTi. It is possible that the follow-up interval was too short to capture significant changes in CRP in our sample. Notably, studies reporting reductions in CRP [31,32], IL-6 [47], and monocytic production of IL-6 and TNF-*α*, or TNF-*α* only [32] had markedly longer follow-up intervals (i.e., 16 months) than the present and previous studies. Irwin et al. [32] reported reduced monocytic production of IL-6 and TNF-*α* at 2 months, but not at later follow-ups (4, 7, or 16 months) after CBTi, compared with sleep seminar education or Tai Chi Chih. This pattern may suggest early molecular changes that do not necessarily translate into long-term alterations in inflammatory protein levels. The present findings may be explained by our sample being relatively low-risk (i.e., healthy due to exclusion criteria), limiting the ability to detect meaningful changes in inflammatory markers. Indeed, Carroll et al. [33] observed reductions in biological risk encompassing lipid, glucose and inflammatory markers (including CRP) among older adults following sleep improvement, supporting the potential of CBTi as a tertiary prevention strategy to help manage or mitigate disease progression post-diagnosis.

### 4.3. Strengths and limitations

Key strengths of the current study lie in its design and assessments. The design was a randomized-controlled trial, which is considered the gold standard in examining cause-effect associations between treatment and outcomes [48]. Moreover, participants were clinically diagnosed with insomnia and several cardiovascular and immunological assessments were obtained at multiple times.

Limitations that merit consideration include the relatively limited sample size and its limited diversity, as participants were primarily middle-aged, White, female, and university educated. This may limit generalizability and reduce sensitivity to detect between-group differences. In addition, given changes in inflammatory profiles during the menopausal transition, this may have influenced the results in this predominantly middle-aged female population [49]. This study may have been underpowered due to attrition across assessment time points, particularly for inflammatory biomarkers. Retention issues in prospective studies may reflect the demanding protocol (e.g., overnight sleep laboratory assessment) as well as participants’ misconceptions, anxiety, fear, and/or prior negative experiences with blood draws [50]. In addition, key covariates known to influence these outcomes (e.g. body weight, perceived stress, medication or supplement use, and health behaviors) could not be included in the analyses due to the limited sample size. The predominantly healthy nature of the population further reduced variability in biomarkers levels. Future studies should therefore examine these effects in at-risk or clinically diagnosed cardiovascular populations. Finally, the absence of a healthy good-sleep control group limits interpretation of whether baseline alterations in these physiological markers are specific to chronic insomnia, particularly given the mixed and heterogenous nature of the existing literature.

### 4.4. Perspectives and conclusion

Replication of prospective, randomized-controlled trials are essential and should include larger, representative samples with varying levels of health risk/comorbidity and insomnia subtypes (e.g., objectively short sleepers) to better elucidate the putative impact CBTi has on physiological outcomes across different health statuses. Importantly, while larger sample sizes would increase statistical power, our findings suggest that any group-by-time effects in cardiovascular and inflammatory biomarkers are likely to be small. Accordingly, future studies with larger samples are needed to improve precision and further evaluate their clinical relevance. In addition, incorporating multiple and well-timed follow-up intervals may help better characterize the temporal evolution of intervention effects. Finally, given individual differences to treatment acceptance and efficacy, it is imperative to examine differences between pharmacological and non-pharmacological interventions for insomnia and pathophysiological indices [37].

In conclusion, the present study investigated the potential physiological benefits of CBTi. The published parent study on this same dataset showed an improvement in sleep perception following CBTi, without corresponding changes in objective sleep measures [39]. In our current study, no significant changes were observed in cardiovascular or inflammatory biomarkers. Overall, these findings suggest that physiological responses associated with CBTi may be complex and not captured by conventional systemic biomarkers. While the intervention improves subjective sleep experience, its impact on biological pathways related to cardiovascular and inflammatory regulation remains unclear in the present sample. These results highlight the complexity of physiological adaptations following CBTi and suggest that treatment-related changes may involve more dynamic, context-dependent or sleep stage-related regulatory mechanisms. Given that persistent insomnia is associated with substantial adverse consequences for mental health, cardiometabolic risk and broader societal impacts, further research is warranted to better characterize the biological processes underlying insomnia/CBTi effects and their potential role in early preventive strategies.

## AUTHOR CONTRIBUTIONS

Conceptualization: TTDV, JPG, MR and DC; Methodology: MR and DC. Formal Analysis: MR and DC; Investigation: AAP, FB, DS, KG, LT; Data Curation: MR, DC, AAP, FB, KG, LT; Writing – Original Draft: MR, DC, TTDV and JPG; Writing – Review & Editing: MR, DC, AAP, FB, DS, KG, LT, JS, TTDV and JPG; Visualization: MR; Project Administration: AAP, FB, LT; Funding Acquisition: TTDV, JPG and JS. All authors approved the final version of the manuscript.

## DATA AVAILABILITY STATEMENT

The data presented in this study are available from the corresponding author upon reasonable request due to privacy reasons.

## ACKNOWLEDGEMENTS

We acknowledge the contributions of the following students who assisted in participants’ recruitment, data collection and data preprocessing: Jennifer Suliteanu, Kazem Habibi, Brian Hodhod, Alex Hillcoat, Kajamathy Subramaniam, Elizaveta Frolova, Emma-Maria Phillips, Rachel Hu, Loren Bies, Meaghan Pawlowski, Aminata Balde, Alexandros Hadjinicolaou, Elissa Pierre, Victoria Yue, Laurence Vo Buu, Maryam Aboutiman, Shira Azoulay and all the volunteers. We acknowledge the contribution of the psychologists who provided CBTi, the research coordinators, and nurses. We also thank our sleep technologists Madeline Dickson and Elinah Mozhentiy, and Liza Perez from the Clinique SomnoMed for their contribution to the setup of sleep recordings. Finally, we would like to thank the participants for giving their time and energy into this research study.

