## Supplementary materials for "The Effects of Cognitive Behavioral Therapy for Insomnia on Cardiovascular and Immunological Outcomes: A Randomized-Controlled Study"

Department of Psychology,
Concordia University
7141 Sherbrooke Street West, Montréal, QC H4B 1R6, Canada

SUPPLEMENTARY METHODS

### Participant retention and missing data

**Blood Pressure.** At baseline (T1), systolic (SBP) and diastolic blood pressure (DBP) data were available for 31 of 33 participants (93.9%) in the TX group. At post-intervention (T2), blood pressure data were provided by 29 of 33 participants (87.9%) in the TX group and 22 of 29 participants (75.9%) in the WL group. There were no differences in age or sex between groups at baseline ($p_s$>0.05). There were no significant differences in age or sex between participants who did and did not provide T2 measures in either the TX or WL group ($p_s$>0.05).

For the pooled data, of the 62 participants included at baseline, 54 (87.1%) provided post-CBTi blood pressure measurements. There were no differences in age or sex between participants who provided or did not provide post-treatment assessments ($p_s$>0.05).

**Heart Rate (HR).** At baseline, HR data were available for 31 of 33 participants (93.9%) in the TX group and 28 of 29 participants (96.6%) in the WL group. At T2, HR measurements were obtained for 29 of 33 participants (87.9%) in the TX group and 21 of 29 participants (72.4%) in the WL group. No differences were observed in age or sex between groups at baseline ($p_s$>0.05). There were no significant differences in age or sex between participants who did and did not provide T2 measures in either the TX or WL group ($p_s$>0.05).

For the pooled data, post-CBTi HR data were provided by 47 of 62 participants (75.8%). There were no differences in age or sex between participants who provided or did not provide post-treatment heart rate measurements ($p_s$>0.05).

**Heart Rate Variability (HRV).** At T1, data were collected from 32 of 33 participants (97.0%) in the TX group and all 29 participants (100%) in the WL group. At T2, 26 of 33 participants (78.8%) in the TX group and 22 of 29 participants (75.9%) in the WL group completed the HRV assessment. No differences in age or sex between groups at baseline ($p_s$>0.05), nor between participants who did and did not provide T2 measures within the TX and WL groups ($p_s$>0.05).

For the pooled data, post-CBTi measurements were available for 47 of 62 participants (75.8%). No differences in age or sex were observed between participants who did and did not provide post-treatment measures within either the TX or WL group ($p_s$>0.05). There was no difference in sex between participants who provided or did not provide post-treatment assessments ($p_s$>0.05). However, participants who did not provide T2 measures ($M$=43.3 years, $SD$=16.55) were younger than those who did ($M$=52.6 years, $SD$=14.79) with a trend toward significance ($t$=2.06, $p$=.05).

**Inflammatory markers.** At T1, blood samples were collected for 32 of 33 participants (97.0%) in the TX group and for all 29 participants (100%) in the WL group. At T2, measurements were completed by 29 of 33 participants (87.9%) in the TX group and 23 of 29 participants (79.3%) in the WL group. No differences in age or sex between groups at baseline ($p_s$>0.05), nor between participants who did and did not provide T2 measures within the TX and WL groups ($p_s$>0.05).

For the pooled data, 48 of 62 participants (77.4%) contributed post-CBTi inflammatory marker measurements. No differences in age or sex were observed between participants who did and did not provide post-treatment assessments ($p_s$>0.05).

SUPPLEMENTARY RESULTS

**Effect sizes of the effects of CBTi compared to a waitlist control group on inflammatory and cardiovascular outcomes**

Effect sizes (Hedges' *g*) were derived from estimated marginal means using the residual standard deviation of the mixed-effects model and corrected for small-sample bias. Positive values indicate an improvement in the outcome. Effect sizes were interpreted according to conventional thresholds: 0.2 = small, 0.5 = medium, and 0.8 = large. Confidence intervals were reported alongside Hedges' *g* to provide an estimate of precision.

**Table S1.** Between-group and within-group effect sizes with 95% confidence intervals

| | Between-group difference at T1 | Between-group difference at T2 | Change in group WL (T2 vs T1) | Change in group TX (T2 vs T1) |
|---|---|---|---|---|
| SBP (mmHg) | 0.47 [-0.40 – 1.35] | 0.77 [-0.16 – 1.71] | -0.17 [-0.76 – 0.42] | 0.13 [-0.16 – 1.71] |
| DBP (mmHg) | 1.10 [0.02 – 2.21] | 1.30 [0.17 – 2.46] | 0.11 [-0.48 – 0.71] | 0.31 [-0.22 – 0.85] |
| HR (bpm) | -0.05 [-0.93 – 0.84] | 0.38 [-0.56 – 1.34] | -0.31 [-0.94 – 0.32] | 0.12 [-0.41 – 0.65] |
| SDNN (ms) | -0.68 [-2.14 – 0.78] | -0.85 [-2.38 – 0.66] | 0.26 [-0.36 – 0.87] | 0.08 [-0.52 – 0.68] |
| RMSSD (ms) | -0.47 [-1.71 – 0.76] | -0.44 [-1.74 – 0.85] | 0.19 [-0.41 – 0.79] | 0.22 [-0.37 – 0.82] |
| Log HF | -0.44 [-1.72 – 0.83] | -0.31 [-1.64 – 1.02] | 0.08 [-0.52 – 0.68] | 0.21 [-0.38 – 0.81] |
| Log LF | -0.57 [-1.95 – 0.79] | -0.79 [-2.23 – 0.63] | 0.28 [-0.34 – 0.89] | 0.05 [-0.54 – 0.65] |
| SDNN (ms) 90 minutes | -0.25 [-1.49 – 0.98] | -0.43 [-1.73 – 0.86] | 0.02 [-0.58 – 0.62] | -0.16 [-0.76 – 0.44] |
| RMSSD (ms) 90 minutes | -0.36 [-1.69 – 0.96] | -0.57 [-1.96 – 0.81] | 0.32 [-0.28 – 0.92] | 0.11 [-0.48 – 0.71] |
| Log HF 90 minutes | -0.21 [-1.56 – 1.14] | -0.30 [-1.71 – 1.10] | 0.22 [-0.37 – 0.83] | 0.13 [-0.47 – 0.73] |
| Log LF 90 minutes | -0.31 [-1.46 – 0.84] | -0.37 [-1.59 – 0.84] | -0.04 [-0.64 – 0.55] | -0.11 [-0.70 – 0.49] |
| Log CRP | 0.92 [-0.47 – 2.31] | 1.04 [-0.38 – 2.47] | -0.09 [-0.68 – 0.49] | 0.03 [-0.50 – 0.56] |
| Log TNF-$\alpha$ | 0.66 [-0.45 – 1.78] | 0.85 [-0.31 – 2.02] | -0.16 [-0.76 – 0.44] | 0.03 [-0.50 – 0.57] |
| Log IL-6 | 0.48 [-0.48 – 1.46] | 0.23 [-0.79 – 1.25] | 0.01 [-0.57 – 0.59] | -0.24 [-0.77 – 0.29] |
| BDNF | 0.56 [-0.25 – 1.38] | 0.15 [-0.72 – 1.03] | 0.21 [-0.38 – 0.80] | -0.20 [-0.73 – 0.33] |

***Note.*** WL = Waitlist; TX =Treatment; SBP = Systolic blood pressure; DBP = Diastolic blood pressure; HR = Heart rate; SDNN = standard deviation of normal-to-normal intervals; RMSSD = root mean square of successive differences; HF = high frequency; LF = low frequency; CRP = C-reactive protein, TNF-$\alpha$ = Tumor necrosis factor-alpha, IL-6 = Interleukin-6. BDNF = Brain-Derived Neurotrophic factor.

**Descriptive statistics and effect sizes for cardiovascular and immunological variables following CBTi exposure in both groups pooled**

Effect sizes (Hedges' g) were computed as model-based standardized mean differences derived from estimated marginal means, using the residual standard deviation of the fitted mixed-effects model and corrected for small-sample bias.

**Table S2.** Descriptive Statistics for cardiovascular and immunological variables following CBTi exposure in both groups pooled and effect sizes of time Effects (T2-CBTi, T3-WL)

| | **Pre-treatment** M $\pm$ SD | **Post-treatment** M $\pm$ SD | **Effect size** Hedges' $g_{Time}$ [95% CI] |
|---|---|---|---|
| SBP (mmHg) | 117.13 $\pm$ 13.34 | 115.78 $\pm$ 13.43 | -0.27 [-0.68 – 0.13] |
| DBP (mmHg) | 68.57 $\pm$ 8.61 | 67.97 $\pm$ 8.00 | -0.24 [-0.65 – 0.17] |
| HR (bpm) | 68.55 $\pm$ 9.57 | 68.06 $\pm$ 9.61 | 0.01 [-0.42 – 0.42] |
| SDNN (ms) | 65.69 $\pm$ 18.91 | 64.25 $\pm$ 20.61 | 0.02 [-0.40 – 0.45] |
| RMSSD (ms) | 70.87 $\pm$ 17.87 | 68.12 $\pm$ 17.63 | -0.08 [-0.50 – 0.34] |
| Log HF | 7.27 $\pm$ 0.56 | 7.18 $\pm$ 0.62 | -0.09 [-0.51 – 0.34] |
| Log LF | 7.33 $\pm$ 0.69 | 7.26 $\pm$ 0.77 | -0.01 [-0.43 – 0.42] |
| SDNN (ms) 90 minutes | 59.61 $\pm$ 18.44 | 58.75 $\pm$ 21.75 | 0.05 [-0.37 – 0.47] |
| RMSSD (ms) 90 minutes | 68.58 $\pm$ 17.68 | 65.61 $\pm$ 18.52 | -0.06 [-0.49 – 0.36] |
| Log HF 90 minutes | 7.14 $\pm$ 0.60 | 7.03 $\pm$ 0.67 | -0.13 [-0.56 – 0.29] |
| Log LF 90 minutes | 7.12 $\pm$ 0.75 | 7.05 $\pm$ 0.89 | -0.01 [-0.42 – 0.44] |
| Log CRP | 5.85 $\pm$ 0.55 | 5.87 $\pm$ 0.57 | -0.04 [-0.45 – 0.36] |
| Log TNF-$\alpha$ | 0.09 $\pm$ 0.14 | 0.09 $\pm$ 0.18 | -0.19 [-0.60 – 0.22] |
| Log IL-6 | -0.37 $\pm$ 0.24 | -0.32 $\pm$ 0.25 | 0.32 [-0.08 – 0.72] |
| BDNF | 3.54 $\pm$ 0.23 | 3.52 $\pm$ 0.24 | -0.18 [-0.58 – 0.21] |

***Note.*** M = Mean; SD = Standard deviation; SBP = Systolic blood pressure; DBP = Diastolic blood pressure; HR = Heart rate; SDNN = standard deviation of normal-to-normal intervals; RMSSD = root mean square of successive differences; HF = high frequency; LF = low frequency; CRP = C-reactive protein; TNF- $\alpha$ = Tumor necrosis factor-alpha; IL-6 = Interleukin-6; BDNF = Brain-Derived Neurotrophic factor.
